# *CR1* variants contribute to FSGS susceptibility across multiple populations

**DOI:** 10.1101/2023.11.20.23298462

**Authors:** Rostislav Skitchenko, Zora Modrusan, Alexander Loboda, Jeffrey B. Kopp, Cheryl A. Winkler, Alexey Sergushichev, Namrata Gupta, Christine Stevens, Mark J. Daly, Andrey Shaw, Mykyta Artomov

**Affiliations:** ITMO University, St. Petersburg, Russia; Almazov National Medical Research Centre, St. Petersburg, Russia; Research Biology, Genentech Inc., San Francisco, CA, USA; Broad Institute, Cambridge, MA, USA; Kidney Disease Section, Kidney Diseases Branch, National Institute of Diabetes and Digestive and Kidney Diseases (NIDDK), NIH, Bethesda, Maryland, USA; Molecular Genetic Epidemiology Studies Section, National Cancer Institute (NCI), Frederick, Maryland, USA; Massachusetts General Hospital, Boston, MA, USA; Institute for Molecular Medicine Finland, Helsinki, Finland; Institute for Genomic Medicine, Nationwide Children’s Hospital, Columbus, OH, USA; Department of Pediatrics, The Ohio State University College of Medicine, Columbus, OH, USA

## Abstract

Focal segmental glomerulosclerosis (FSGS) is a common cause of nephrotic syndrome with an annual incidence in the United States in African-Americans compared to European-Americans of 24 cases and 5 cases per million, respectively. Among glomerular diseases in Europe and Latin-America, FSGS was the second most frequent diagnosis, and in Asia the fifth. We expand previous efforts in understanding genetics of FSGS by performing a case-control study involving ethnically-diverse groups FSGS cases (726) and a pool of controls (13,994), using panel sequencing of approximately 2,500 podocyte-expressed genes. Through rare variant association tests, we replicated known risk genes – *KANK1*, *COL4A4,* and *APOL1*. A novel significant association was observed for the gene encoding complement receptor 1 (*CR1)*. High-risk rare variants in *CR1* in the European-American cohort were commonly observed in Latin-and African-Americans. Therefore, a combined rare and common variant analysis was used to replicate the *CR1* association in non-European populations. The *CR1* risk variant, rs17047661, gives rise to the Sl1/Sl2 (R1601G) allele that was previously associated with protection against cerebral malaria. Pleiotropic effects of rs17047661 may explain the difference in allele frequencies across continental ancestries and suggest a possible role for genetically-driven alterations of adaptive immunity in the pathogenesis of FSGS.

## Introduction

Focal segmental glomerulosclerosis (FSGS) is a common cause of primary nephrotic syndrome among both adults and children in the USA, and its incidence is increasing.^1,2^ The incidence and prevalence of FSGS are not precisely known due to the requirement of a kidney biopsy for diagnosis and the lack of a central registry. Estimates for incidence range from 1.4 to 21 cases per million population.^3^ The incidence of FSGS in the U.S. is about 4 times higher in African Americans (6.8 patients per million) and 2 times higher in Latin-Americans (3.7 patients per million) compared to European-Americans (1.9 patients per million).^4^

Genetic studies of FSGS, conducted using both pedigree analyses and cohort-based association studies, have identified a number of susceptibility genes, yet explaining only a fraction of family-history enriched cases.^5^ The first genetic studies identifying the chromosome (chr) 22 region with FSGS were prompted by the observed higher prevalence of FSGS in African and African-American populations, suggesting that one or more FSGS susceptibility gene variants would be enriched on African-derived haplotypes.^6–8^ Thesubsequentdiscoveryof association of G1 and G2 coding variants in *APOL1* with FSGS provided an explanation for increased prevalence of the disease among African-descent populations. Pleiotropic properties of these variants resulted in protection against trypanosomiasis but at the cost of increased FSGS risk.^9^

In this study, a large-scale genetic database was assembled from biopsy-confirmed cases of Focal Segmental Glomerulosclerosis (FSGS) and ethnically matched controls. The study used a panel of approximately ∼2,500 genes associated with podocytes, which play a crucial role in the formation and maintenance of the glomerular filtration barrier. The purpose of the study was to investigate the genetic basis of FSGS and identify novel susceptibility genes. This study is a significant extension of previous work conducted by Yu et al^5^, with increased power and a more diverse multi-ethnic cohort with greater sample size.

## Methods

DNA samples were obtained from patients participating in a multicenter NIDDK study of biopsy-confirmed FSGS^6^ and from patients diagnosed at Washington University. The research protocols were approved in advance and all subjects provided informed consent or assent. As all samples were de-identified, the Washington University in St. Louis Institutional Review Board (IRB) deemed that these studies did not require IRB approval. A total of 726 samples were collected in a multicenter NIH study of biopsy-confirmed FSGS^6^ and from patients similarly diagnosed at Washington University, the latter inherited from Yu et al^5^. Genetic data for cases were obtained using a “podocyte exome” sequencing approach, consisting of a panel of 2,482 genes, as described in Yu et al^5^. Of the selected genes, mutations in five genes cause familial FSGS and 200 genes are functionally related to these five. Additional genes were selected based on expression profiles, as 677 genes are highly expressed in human micro-dissected glomeruli, and 1600 genes are human orthologs of highly-expressed mouse podocyte genes (**Figure 1A)**. In the present study using this panel, only the 2,482 genes constituting the “podocyte exome” were analyzed, with 1.12% of the sequencing capture nucleotides located in non-coding DNA.

**Figure 1.**
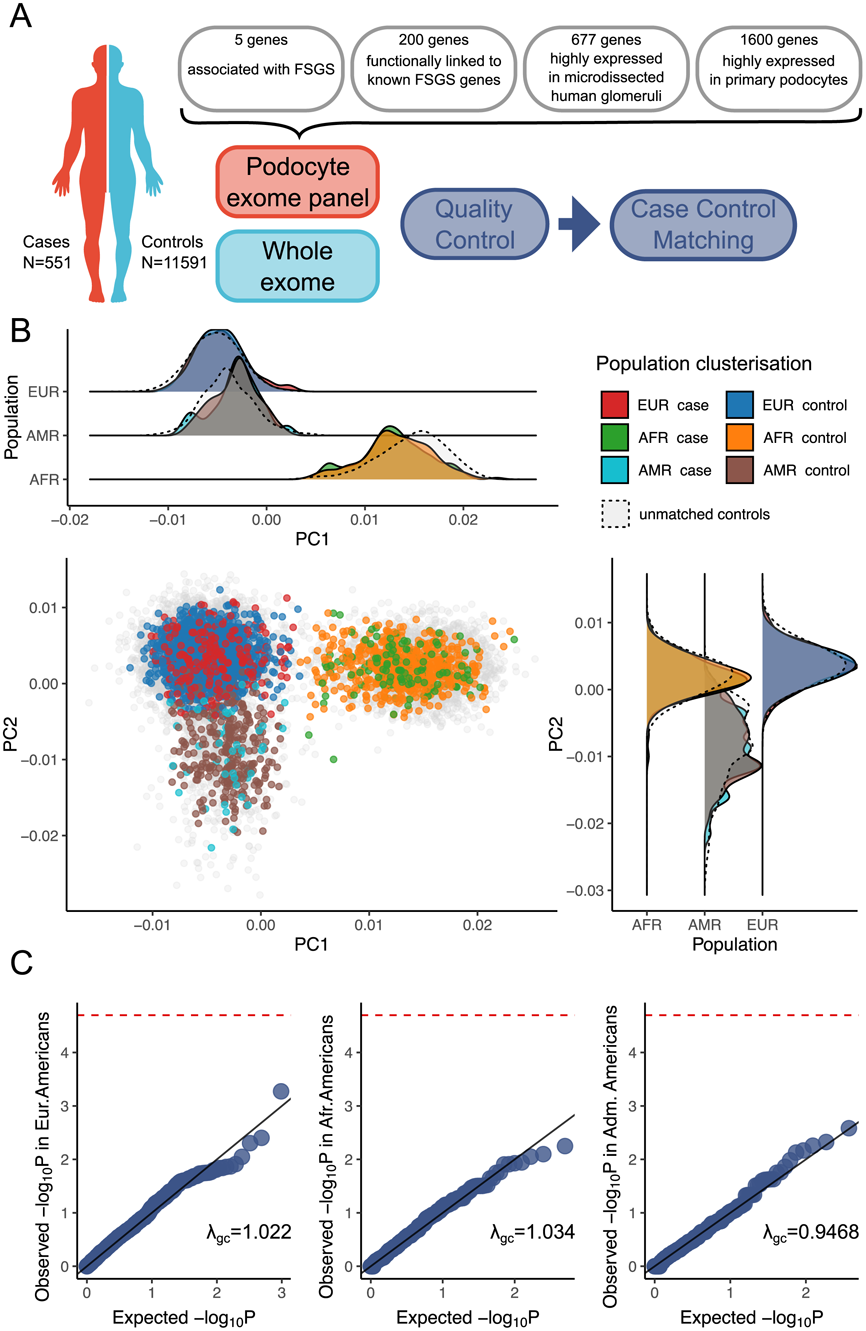
Study design, principal component analysis, and quantile-quantile plots to identify calibration of synonymous variants between case and control cohorts. (A) **Case-control study design**. DNA samples from FSGS cases and controls were examined for coding variants in a podocyte exome panel gene panel composed of 2482 genes and also was subjected to whole exome analysis. (B) **Principal component analysis** illustrates case-control matching in European-derived, African-derived and Latin-American-derived-populations and demonstrates genetic segregation of these three populations. (C) **Quantile-quantile (QQ)-plots for the association study of the common synonymous variants with gnomAD population specific allele frequency ≥0.01).** These plots illustrate case-control matching quality for the European-derived (left), African-derived (middle), Latin-American-derived (right) populations. The test lambda-GC (genomic inflation factor) for genome-wide association studies (GWAS) compares the median test statistic against the expected median test statistic under the null hypothesis, in which there is no association for each variant. This test identifies systemic biases and significant associations. Here, most of the points fall along the diagonal, indicating the absence of systemic bias. Abbreviations. EUR, European-Americans. AFR, African-Americans. AMR, Admixture Americans

The raw data files with sequencing reads (FASTQ files) were obtained for control subjects from dbGAP general population cohorts not ascertained for kidney disease history (**Sup. Table S1)**. We extracted the regions sequenced in the podocyte exome from the full-exome data of the control cohort. There were 333,239 variants in the raw dataset of 726 cases and 13,994 controls. We performed joint variant calling according to GATK best practices,^10^ to construct a case-control dataset. To confirm the absence of insufficient coverage biases between cases and controls, we crossed the intervals common to both panels and then we calculated the fraction of sequencing intervals that were well-covered (>10X) in cases and controls; this was found to be 89% for both groups (**Figure S1)**. In this calculation, only variants that passed initial GATK hard-filtering^10^ were used. Next, the case-control dataset was subjected to quality filtration using the Hail 0.2 open source software library^11^ (**Figure S2)**.

The final data set included 577 cases (including 179 from Yu et al^5^), (including 378 from Yu et al^5^), and 131,179 variants. The drop-out rate was 60.64% for variants and 6.85% for samples. The high drop-out rate for variants is explained by exome sequencing being joined with panel sequencing in a single dataset, requiring exclusion of many variants detected in the exome sequences of controls and not sequenced in the case panel, due to broader DNA region coverage in controls.

To account for population stratification, a joint principal components analysis (PCA) was performed for case and control genotypes. Uncorrelated common variants (linkage disequilibrium pruning: r^2^<0.9; minorallelefrequency– MAF>0.05) wereusedtoclusterthe samples in PCA space. To reduce the risk of false associations in the rare variant analysis due to population stratification, we used principal components to subset the control cohort to match the genetic background of cases.

First, we partitioned the dataset into clusters representing global population groups. Clustering was performed using mixed Gaussian models AutoGMM package^12^. The data were stratified into eight clusters according to the Gaussian mixture model. Agnostic clusters modeled by the AutoGMM algorithm were mapped to known clusters of the 1000 Genomes Project and labeled accordingly **(Figure S3)**. Two minor clusters of individuals belonging to South Asian and East Asian populations and admixed ancestry were excluded from the analysis due to small case count in each cluster, resulting in low statistical power. Of the six clusters retained for further study, three included individuals of Europe and escent and reflected different local-population origins; these minor-clusters were combined into a single major-European cluster. The fourth cluster represented the individuals with African ancestry. Two other clusters belonged to the Latin-American population. After filtering out the low power clusters, the dataset had 551 cases and 11,591 controls (**Figure S4)**.

Further case-control matching was conducted using the MatchIt^13^ package (**Figure 1B, Figure S4)**. 16 cases were excluded from further consideration because it was not possible to select appropriate population controls for them. The final dataset included 358 cases and 1,488 controls for the European-American cluster, 125 cases and 137 controls for the African-American cluster and 52 cases and 288 controls for the Latin-American cluster **(Figure S4)**. The Weir and Cockerham F-statistic for analysis of population structure showed that the European-American cluster and the African-American cluster were sufficiently isolated from each other (weighted mean fixation index Fst=0.0878 for case cohorts), demonstrating distinct population differences. The Latin-American cluster is also sufficiently isolated from the other clusters (weighted mean fixation index Fst=0.0148). The power analysis for the European-American dataset showed many-fold power superiority over previous FSGS cohort studies^14^, and that an exponential power increase threshold has been reached with the current number of cases, *i.e.*, a significant power increase could not be achieved with a larger number of controls (**Figure S5**). The values of significant power for the African and Latin-American clusters are comparable to those of similar studies on European cohorts, but they may not be sufficient to detect genome-wide significant associations of frequent variants because of their small effect size (**Figure S6)**.

We performed an association study using common synonymous variants (gnomAD population specific AF ≥ 0.01), as these are unlikely to contribute to a phenotype and yet reflect possible ancestral bias between case and matched controls cohorts. For all three European-American, African-American and Latin-American post-matching datasets we confirmed the absence of systematic bias between cases and controls (**Figure 1C)**.

## Results

Using a data set of matched cases and controls after quality filtration, we performed several association studies. Because analyses of this dataset were limited to the “podocyte exome”, we focused on missense variants and protein truncation variants (PTV).

For each cluster separately, we conducted a variant-based association study using linear regression with no additional covariates. 3,777 variants with missense and PTV (stop_gained, frameshift_variant, splice_acceptor_variant, splice_donor_variant) effects on protein were included in this analysis. In the European-American cluster, two variants were significantly associated with FSGS: (1) rs601314 – p=8.1×10^-9^, reference allele is a minor allele; OR_minor allele_ =13.24 (CI _95%_ =[3.996, 56.51]), missense,*EFEMP2*(EGF-containing fibulin extracellular matrix protein 2); and (2) rs117071588 – p=4.0×10^-6^, alternative allele is a minor allele; OR _minor allele_ =11.66 (CI _95%_ =[2.790, 68.35]), missense, *CCDC82* (coiled-coiled domain containing 82), Significance threshold was determined with Bonferroni correction - p<0.05/3,777=1.32×10^-5^; **Figure S7**, **Sup. Table S2)**. Replication analysis of these two variants in the African-American cohort showed that the rs601314 variant (*EFFMP2)* was not significantly associated with FSGS (p=0.062, reference allele is a minor allele, OR_minor allele_ =0.7418, CI _95%_ =[0.5370, 1.015]) and that rs117071588 (*CCDC82)* was absent from the African-American dataset. A similar situation was observed when replication was attempted in the Latin-American cohort: rs601314 was not significantly associated (p=0.097, reference allele is a minor allele, OR_minor allele_ =2.99, CI_95%_ =[0.6891, 14.76]), rs117071588 was absent from the data. Despite the significant association statistics of *EFEMP2* and *CCDC82* in the European-American group, the lack of replication of the *EFEMP2* variant FSGS association in the other populations makes this a less robust finding but might serve as a starting point for future studies.

The rs601314 in *EFEMP2* (NC_000011.9:g.65636053T>C, ENSP00000434151:p.I259V) and rs117071588 in *CCDC82* (NC_000011.9:g.96117537A>C, ENSP00000278520:p.D125E) variants are defined by most common *in silico* pathogenicity predictors as benign^15^. FATHMM classified rs601314 as “damaging” (Fathmm Score Converted = 0.48)^15^. The specific predictors of missense deleteriousness classified rs601314 and rs117071588 as benign (MISTIC<0.5)^16^. Variant rs601314 affects the von Willebrand factor type A (vWA) domain of *EFFMP2* and variant rs117071588 affects the domain of unknown function (DUF4196) of *CCDC82*^15^. Both variants have a missense effect on protein function and come from non conservative parts of proteins (MPC<2.0)^17^.

We carried out rare variant burden analyses in the European-American cohort, focusing on missense variants and PTV with a population frequency below 0.01. This cutoff was chosen according to the presence of a signal in each of the quartiles of allele frequency distribution in the interval [0; 0.01] (**Figure S8)**. A rare variant association study (RVAS) was performed using five tests representing different statistical classes of methods for each gene, in order to identify all potential risk patterns. If most variants are causal and have unidirectional effects, classical burden tests are useful, due to their high power. However, adaptive burden tests are considered more reliable than those using fixed weights or thresholds.^18^ In addition, some tests can improve understanding of the results. Tests of variance components are effective when there are variants that both increase and decrease a trait, or have a limited number of causal variants^18^. These tests included Fisher’s exact test, C-alpha, adaptive sum statistic (ASUM), weighted sum statistics (WSS) and kernel-based adaptive clustering (KBAC) (**Figure S9)**. The resulting p-values were combined using the Simes method for multiple hypothesis testing, which is suitable for merging dependent test statistics (**Sup. Table S3)**. The top 10 associated genes included four genes, *APOL1*, *KANK1*, *COL4A4*, *IL36G,* that were previously identified in FSGS association studies. Of these, the top two genes reached significance after Bonferroni correction (p=0.05/2,482=2.015×10^-5^): *APOL1* (p=1.47×10^-6^), a known FSGS susceptibility gene, and *CR1* (p=1.67×10^-5^), a novel candidate gene (**Figure 2A, Sup. Figure S10)**.

**Figure 2.**
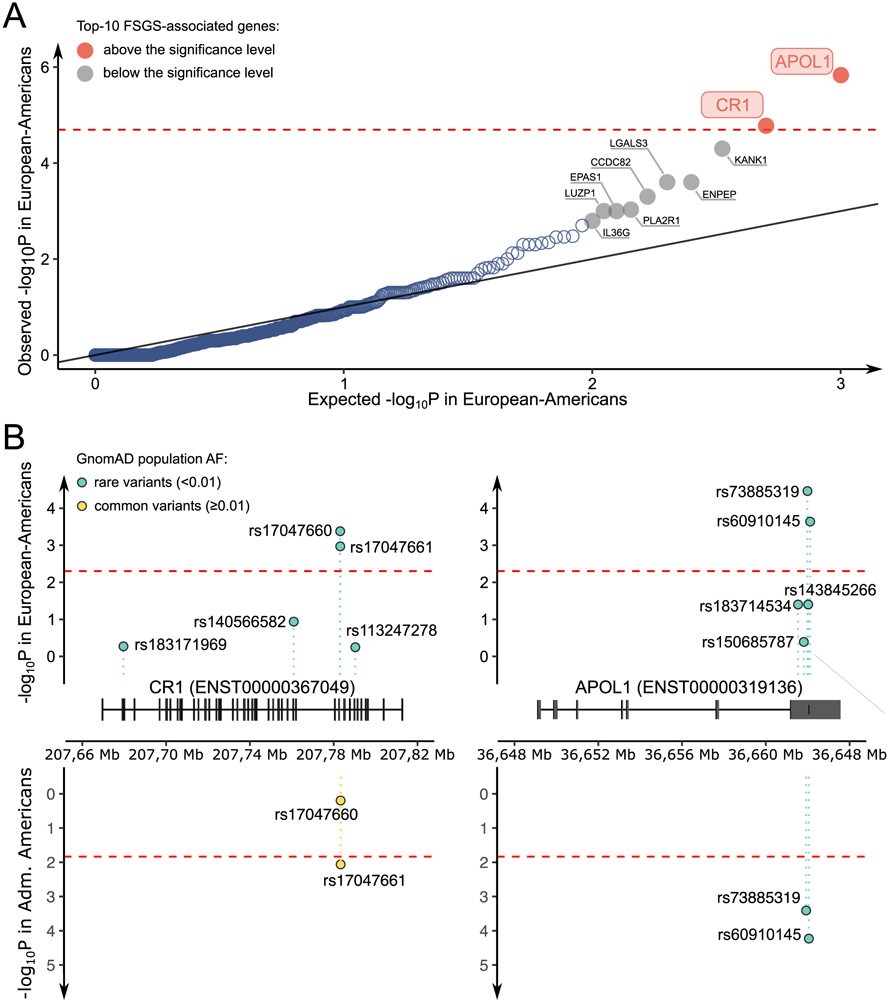
Rare variant association study in the European-American cohort and replication of *CR1* and *APOL1* variants in the Admixed American cohort. **A)** Shown graphically are the results of rare-variant association study involving the European-American subject cluster (gnomAD EUR AF < 0.01; missense and PTV (protein truncating variants). Statistical approaches included the following: the Simes method for multiple hypothesis testing, the Fisher exact test for testing two groups, C-alpha test for comparing the variance of each group against the expected, adaptive sum statistic (ASUM) for testing variants;, weighted sum statistics (WSS) for testing variants and kernel-based adaptive clustering (KBAC) for variant classification and association testing. Of the top 10 (most significant) genes, four with known FSGS-associated variants. *CR1*, complement C3b/C4B receptor 1; *KANK1*, KN motif and ankyrin repeat domains 1; *COL4A4*, collagen type 4, alpha 4 chain; IL-36G, interleukin 36G. **(B)** Shown are associations of FSGS-related SNPs in *CR1* and *APOL1* in the GnomAD aggregation database population among European-Americans (top panels) and Admixed-Americans (bottom panels). **Left Panels.** Two rare variants in *CR1* have been associated with FSGS in Euro-Americans, and one of these variants in *CR1* has also been associated with FSGS in Latin-Americans (both variants in *CR1* are common in Latin-Americans) **. Right panels**. Two rare variants in *APOL1* are associated with FSGS in European-Americans and in Latin-Americans.

Significantly-associated genes in the European-American cohort were further examined in a replication study of the African-American and Latin-American cohorts. Neither *APOL1* nor *CR1* were replicated using rare (MAF<0.01) variant analysis (**Sup. Table S4)**. Previously-observed positive selection acting on the *APOL* ^9^ variants in the African-American population suggests that FSGS risk variants might be too common to be detected by a RVAS in non-European populations. Therefore, we used the variant-based tests to replicate the FSGS- association signals in *APOL1* and *CR1*.

We identified rare variants in the European-American cohort that drove the association signals in *APOL1* and *CR1* and four variants, consisting of pair-locus G1 in *APOL1* (rs60910145 [NC_000022.10:g.36662034T>G, ENSP00000317674.4:p.Ile400Met] and rs73885319 [NC_000022.10:g.36661906A>G, ENSP00000317674.4:p.Ser358Gly]^9^) and two closely-adjacent variants in *CR1* (rs17047661 [NC_000001.10:g.207782889A>G, ENSP00000356016.4:p.Arg2051Gly] and rs17047660 [NC_000001.10:g.207782856A>G, ENSP00000356016.4:p.Lys2040Glu]) were selected for replication in other ancestries (**Figure 2B)**. Additionally, we eliminated the possibility of this result being a false positive due to coverage imbalance in the associated genes (**Sup. Figure S11)**. *APOL1* variants were successfully replicated (rs73885319: p=0.001779, alternative allele is a minor allele, OR_minor allele_ =1.59, CI _95%_ =[1.18, 2.14]; rs60910145: p=0.002271, alternative allele is a minor allele, OR _minor allele_ =1.58, CI _95%_ =[1.17, 2.12]). The variants in *CR1* were more common and had smaller effect size, therefore, we lacked the statistical power to see the significant replication (rs17047661: p=0.28, reference allele is a minor allele, OR _minor allele_ =1.22, CI _95%_ =[0.84, 1.76]; rs17047660: p=0.74, alternative allele is a minor allele, OR _minor allele_ =1.09, CI _95%_ =[0.69, 1.71]).

Second replication was attempted in the Latin-American cohort because the variant frequencies for the variants of interest are more similar to the original European-American cohort. Variants rs17047661 in *CR1* and rs60910145 and rs73885319 in *APOL1* surpassed the replication significance threshold (p=0.05/4=0.0125) (**Sup. Table S5**). Analysis of the statistical power for identified effect sizes in Latin-American and African-American cohorts indicated that the lack of replication in the latter is most likely driven by the statistical power limitations (**Sup. Figure S6)**.

Meta-estimates of pMETAL^18^ for all 4 variants were also calculated for the European-American and Latin-American cohorts: rs60910145 (p _METAL_ =9.706×10^-6^), rs73885319 (p _METAL_ =1.420×10^-4^), rs17047660 (p _METAL_ =0.6359), rs17047661 (p _METAL_ =9.314×10^-3^). Interestingly, the variants in *CR1*: rs17047661 and rs17047660 are linked with only three out of four possible haplotypes observed in African subpopulations in 1000 genomes (AFR:YRI+LWK+GWD+MSL+ESN+ASW+ACB: r^2^=0.15, D’=1; YRI: r^2^=0.15, D’=1; ASW: r^2^=0.18, D’=1; ACB: r^2^=0.13, D’=1) and observed in global Latin-American population (AMR:MXL+PUR+CLM+PEL: r^2^=0.46, D’=1).

Next, the normalized integral haplotype score (iHS) was directly estimated in the discovery cohort for the variants included in the replication analysis. For the African American cohort, selection pressure analysis confirmed positive selection (iHS<-2.0) for the G1 APOL1 alleles: rs73885319 (iHS=-2.16), rs60910145 (iHS=-2.21) and revealed positive selection for rs17047660 (iHS=-2.71) in CR1, whereas no such selection was detected for rs17047661 (iHS=-1.04). The following results were obtained for the Latin American cohort: rs73885319 (iHS=-1.99), rs60910145 (iHS=-2.01), rs17047660 (iHS=-0.142) and rs17047661 iHS=1.13).

Allele frequencies for all variants included in the replication analyses are significantly different between population groups, which can nominally indicate either positive selection or genetic drift. These included the following variants: rs60910145: gnomAD EUR AF=8.6×10^-5^, gnomAD AFR AF=0.23, p=2.2×10^-16^; rs73885319: gnomAD EUR AF=1.1×10^-4^, gnomAD AFR AF=0.23, p=2.2×10^-16^; rs17047661: gnomAD EUR AF=3.0×10^-3^, gnomAD AFR AF=0.62, p=2.2×10^-16^; and rs17047660: gnomAD EUR AF=1.0×10^-3^, gnomAD AFR AF=0.24, p=2.2×10^-16)^. It is likely that iHS estimates can be skewed by complex population structure or demographic variables such as population growth, bottleneck events, and changes in recombination and mutation frequencies. Notably, the allele frequencies within African and African-American populations in 1000 genomes significantly vary for rs17047661 (AF _YRI_ =0.69, AF _LWK_ =0.70, AF _GWD_=0.79, AF _MSL_=0.79, AF _ESN_ =0.72, AF _ASW_=0.58, AF _ACB_=0.66).

We sought evidence of co-evolving changes in allele frequencies between (a) the G1 and G2 variants in *APOL1* and (b) replicated rs17047661 in *CR1*. We estimated the number of individuals who carry both rs17047661 and either one or both G1 and G2 alleles in the African-American case cohort and compared this with the expectation of random assortment. There were no signs of linkage between these variants (p=0.93, binomial test), which suggests that the effects of rs17047661 are fully independent of those of *APOL1*.

Most common in silico variant effect predictors do not categorize rs17047661 (NC_000001.10:g.207782889A>G, ENSP00000383744:p.R1601G) as pathogenic^15^. Exceptions are, for example, PolyPhen2 HVAR and MutationAssessor, which classify rs17047661 as “probably damaging” (Polyphen 2 Hvar Score = 0.964) and “medium functional effect” (Mutationassessor Score Converted = 0.70), respectively^15^. The rs17047661 variant of CR1 induces a missense effect (p.R1601G) in the protein domains common to secreted complement fixation protein (PHA02927) and complement control protein (CCP) modules, also known as consensus short repeats (SCR) or SUSHI repeats. Specifically, it affects the one of the four Long Homologous Repeats (LHRs), LHR-D, which is responsible for binding C1q, Mannose Binding lectin (MBL) and ficolin^15,19^. However, the specific predictors of missense deleteriousness classified rs17047661 as benign (MISTIC<0.5)^16^, which is likely due to the nonconservative nature of the affected region (MPC<2.0)^17^.

## Discussion

The complement system is a complex network of proteins that play an important role in protecting the body against microbial infections, which are activated either through the classical immune pathway in response to binding to Fc-fragments of IgM or IgG, or through an alternative pathway of non-specific binding to antigens on membranes or to mannose residues through the lectin pathway.^20^ Each protein in the cascade is activated by proteolysis, splitting the original proenzyme into “a” and “b” structures (the exception is C1, which splits into q, r, s molecules). The large molecule “b” is directly involved in the sequential activation of the complement system and the small molecule “a” is an anaphylatoxin, which causes degranulation of mast cells and chemotaxis of other immune cells such as neutrophils, eosinophils, monocytes, and T lymphocytes. All of these factors have the potential to contribute to either innate immune function sort issue injury. C3 is the central element of the complement system, which is activated by C3-convertase, a complex composed of the preceding elements of the cascade (classical/lectin pathway: C4bC2b complex, alternative pathway: C3bBb complex). Upon activation, the complement system can affect cells in two ways: (1) by forming the membrane attack complex (MAC, sС5b-9 complex), resulting in osmotic lysis of the targeted cell, and (2) indirect opsonization through the deposition of C3b on the surface of microbes, which facilitates phagocytosis by immune cells.

*CR1* acts as a negative complement regulator, reducing C3 activation and tissue deposition, by processing and bounding immune complexes, which then facilitates their transfer to the liver or spleen where macrophages ingest and eliminate them. In both the classical and lectin pathways, *CR1* has decay-activating activity, in that its binding C4b prevents the formation of C3-convertase. In the alternative pathway, *CR1* then acts as a cofactor for the cleavage of active C3b fragments (on C3c and C3dg fragments), significantly reducing deposition of C3b fragments, which could activate C3.^21^ CR1 significantly reduces C3b deposition by ∼80% over the classical pathway, but *CR1* is most potent when the alternative pathway is activated (> 95% reduction in C3b deposition).^21^ By stopping the activation of the complement system at the stage of C3-convertase formation, *CR1* is also indirectly involved in reducing the deposition of sC5b-9, which would have formed afterwards if the complement system had been subsequently activated.

*CR1* is expressed by several cell types, including red blood cells (RBC), leukocytes, and among specialized renal cells, *CR1* is localized exclusively on glomerular podocytes. The FSGS related variants in *CR1* – rs17047661 which was replicated in Latin-American descent individuals and its pair – rs17047660 are known as Knops group polymorphisms and are a part of the Red Cell Surface Antigens, which give rise to the Sl2 and McC ^b^ alleles in the Swain-Langley 1 and 2 allele pairs (Sl1/Sl2) and McCoy a and b (McC ^a^/McC^b^), respectively.^22^. The K1590E substitution and the R1601G substitution in *CR1* are located just 11 amino acids away from each other and are in strong LD for both the 1000 genomes for the global African population (r^2^=0.15, D’=1) and for African-American descent from study data (r^2^=0.24, D’=1). They are located in the Long Homologous Repeats (LHRs), motif D, which is responsible for C1q and MBL binding.^22^

No genetic alterations in the adaptive immune system have been identified in FSGS patients to date, and the role of immune and complement systems genetic variants remains unknown. Recent studies have described the role of the complement system in various glomerulopathies.^23^ Typically, the reduction in *CR1* expression is linked to the severity of the disease, as indicated by the degree of inflammation or tissue damage.^24^ Autoantibodies directed against kidney-expressed autoantigens or antibody/antigen complexes deposited in the kidney are causative agents of various human kidney diseases. There are cases of C3-mediated inflammation and deposition.^25,26^ Further, inhibition of C3 reduces proteinuria in animal models.^27^ With regard to FSGS, IgG and C3 deposits are often observed in the affected glomeruli, but the pathogenic role of these deposits remains still unclear, and therapy against the complement system has not been studied in FSGS.^28^

Another study has demonstrated elevated levels of Ba, Bb, C4a, and sC5b-9 in the plasma and urine of patients afflicted with primary FSGS. The detection of these protein deposits in the blood signifies that the complement cascade is activated at a site where fragments can gain entry into the vascular space, likely in the mesangial and sclerotic regions. Conversely, the rise in urine Ba, C4a, and sC5b-9 levels in some patients may reflect complement activation in the glomerulus, or alternatively, activation of filtered proteins in the tubular lumen or downstream in the urinary collection system.^29^ While C5b-9 complexes generally form directly on the membranes of microorganisms, particularly Gram-negative ones, they can also affect adjacent cells, resulting in “bystander” harm.

The relationship between activation of the classical and alternative pathways in response to the presence of Knops antigens in *CR1* has been investigated previously, and a lack of correlation has been noted. However, there is a reasonable discrepancy between the results of serological studies of human samples and those obtained from parts of recombinant proteins. Prior investigations have challenged the conjecture that Sl2 and McC^b^ influence the phenotype by modulating the activity of the cofactor implicated in the cleavage of C3b and C4b or the C1q binding activity.^30^ Nevertheless, these findings warrant future exploration of the involvement of the lectin pathway in the activation of the complement system.^31^ For example, the contribution of the lectin pathway in the activation of the complement system may play a crucial role in the development of progressive glomerular damage and long-term urinary abnormalities in patients with Henoch-Schönlein purpura nephritis (HSPN).^32^ In the case of FSGS, the presence of MBL deposits that are focal and segmental has been observed, as reported in previous studies, and this can also result in tissue damage, MBL deficiency can lead to autoimmune diseases.^33,34^

There are many different pathogens that use *CR1*asareceptorforcellentry,for example: *Leishmania major*^35^, *Legionella pneumophila*^36^, *Leishmania panamensis*^37^ and *Mycobacterium tuberculosis*^38^. It also has been shown that *CR1* is a RBC receptor used by *Plasmodium falciparum* for cell invasion, independent of sialic acid.^39^ Consistently with this hypothesis, Sl2 (rs17047661) was previously associated with protection against cerebral malaria in sub-Saharan African populations, which resulted in much higher prevalence of Sl2 in African-descent individuals compared to Europeans.^19^ In the study conducted by Opi et al, it was noted by the authors that the opposite concomitant effect of the McC ^b^ (rs17047660) allele on the development of severe malaria was of only nominal borderline significance, despite being under significant strong positive selection (iHS<-2.0)^19^. Atfirstglance, theassociation with severe malaria and significant positive McC ^b^ selection are discouraging, but this may explain the linkage of negative and protection haplotypes to each other. Moreover, the authors of the original article tested some haplotypes of Sl and McC combinations and found that the combination of Sl2/McC ^a^ alleles has an additive protective effect against malaria, which may explain the lack of replication signal for rs17047660 in the African-American descent.^19^

In conclusion, the findings reported here establish *CR1* as a novel susceptibility gene for FSGS, involving an autoimmune disease component. Significant alterations in allele frequencies among populations suggest that environmental factors that induce selection pressure, might be responsible for an adaptive benefit, at the cost of kidney disease. These results, together with other evidence of the polygenic nature with many potential mechanisms of FSGS, could be used as a motivation for future GWAS, which would enhance understanding of the molecular genetic mechanisms underlying the disease.

## Data availability

FSGS cohort allele frequencies and gene burden rare allele counts are available in the Supplementary Tables. Raw sequencing data for control cohort subjects are available through the dbGAP, accession numbers are available in the Supplementary Table 1.

## Author contributions

R.S., Z.M., J.B.K., C.A.W., M.J.D., A.S., M.A. designed and conceived the study.

R.S., Z.M., A.L., J.B.K., C.A.W., Al.S., A.S., M.J.D., M.A. analyzed the data

J.B.K., C.A.W., M.J.D., A.S., M.A. acquired funding

N.G., C.S. managed control cohorts

M.J.D., A.S., M.A. supervised the study

R.S., J.B.K., C.A.W., M.J.D., A.S., M.A. wrote the manuscript

All authors reviewed and approved the manuscript

## Acknowledgments

This work was supported in part by the Intramural Research Program, NIDDK, NIH (JBK). This project has been funded in part with federal funds from the National Cancer Institute, National Institutes of Health, under contract 75N91019D00024. The content of this publication does not necessarily reflect the views or policies of the Department of Health and Human Services, nor does mention of trade names, commercial products, or organizations imply endorsement by the U.S. Government. This Research was supported [in part] by the Intramural Research Program of the NIH, National Cancer Institute, Center for Cancer Research and NIDDK. The authors acknowledge the contributions of the following investigators who recruited subjects for FSGS genetic studies, published in Kopp et al, Nature Genetics, 2008, as DNA from those subjects was used in the present study. These investigators include Kopp JB, Freedman BI, Ahuja TS, Berns JS, Briggs W, Cho ME, Dart RA, Kimmel PL, Korbet SM, Michel DM, Mokrzycki MH, Schelling JR, Simon E, Trachtman H. R.S.,Al.S.weresupportedbytheMinistryofScienceandHigherEducationoftheRussian Federation (Priority 2030 Federal Academic Leadership Program).

A.L.wassupportedbyMinistryofScienceandHigherEducationoftheRussianFederation (Agreement # 075-15-2022-301).

M.A. was in part supported by Nationwide Foundation Pediatric Innovation Fund.

