## Supplementary Materials for "*CR1* variants contribute to FSGS susceptibility across multiple populations"

Rostislav Skitchenko[1,2], Zora Modrusan[3], Alexander Loboda[1,2,4], Jeffrey B. Kopp[5], Cheryl A. Winkler[6], Alexey Sergushichev[1], Namrata Gupta[4], Christine Stevens[4], Mark J. Daly[4,7,8], Andrey Shaw[3,#], Mykyta Artomov[4,9,10,#]

[1] – ITMO University, St. Petersburg, Russia
[2] – Almazov National Medical Research Centre, St. Petersburg, Russia
[3] – Research Biology, Genentech Inc., San Francisco, CA, USA
[4] – Broad Institute, Cambridge, MA, USA
[5] – Kidney Disease Section, Kidney Diseases Branch, National Institute of Diabetes and Digestive and Kidney Diseases (NIDDK), NIH, Bethesda, Maryland, USA
[6] – Molecular Genetic Epidemiology Studies Section, National Cancer Institute (NCI), Frederick, Maryland, USA
[7] – Massachusetts General Hospital, Boston, MA, USA
[8] – Institute for Molecular Medicine Finland, Helsinki, Finland
[9] – Institute for Genomic Medicine, Nationwide Children's Hospital, Columbus, OH, USA
[10] – College of Medicine, Ohio State University, Columbus, OH, USA

**A conflict of interest statement is at the end of the manuscript.**

**Supplementary Materials**


**Proportion of covered intervals between cases and controls** 3

**Sequencing data quality filtering** 4

**Case-Control Matching** 6

**Statistical power analysis** 8

**Association studies** Error! Bookmark not defined.

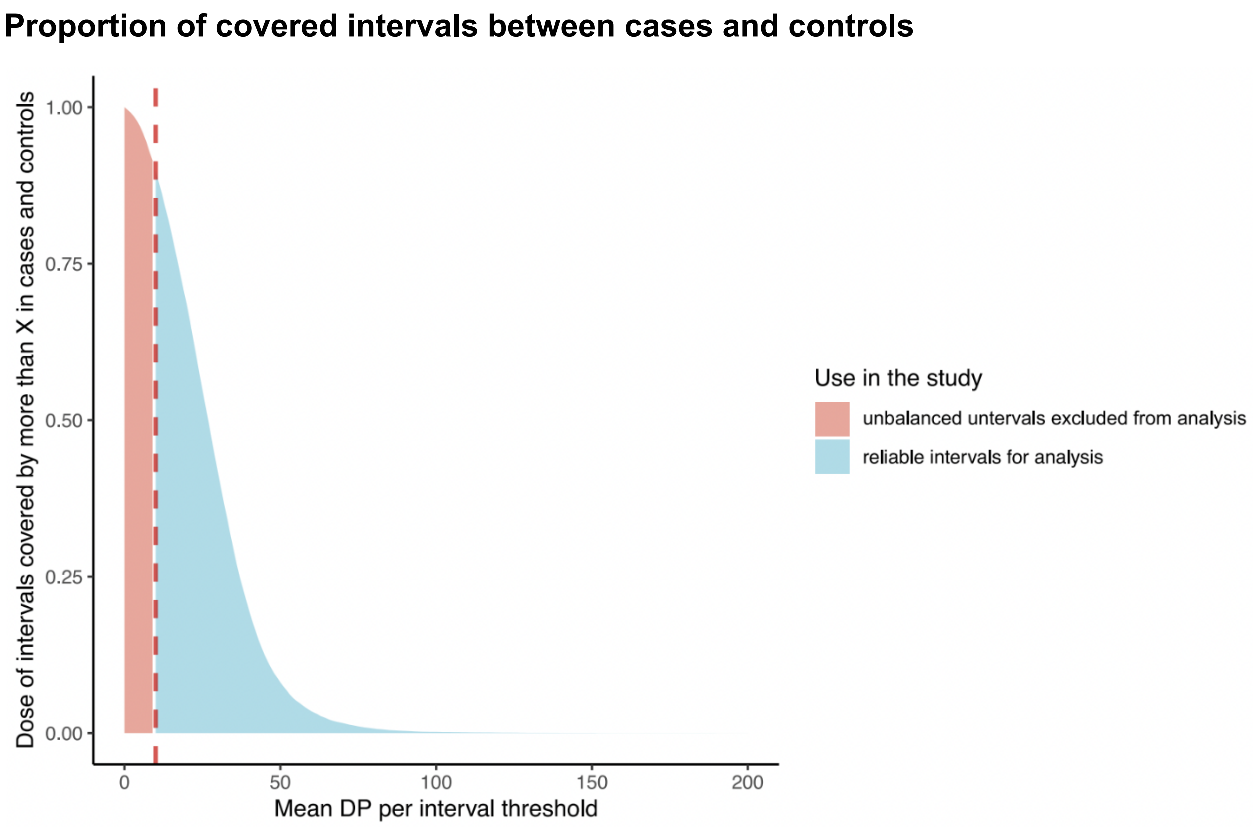


**Figure S1. Proportion of sequenced intervals that were well covered (>10X) in both cases and controls. Only variants that underwent initial hard GATK filtering were used in this calculation.**

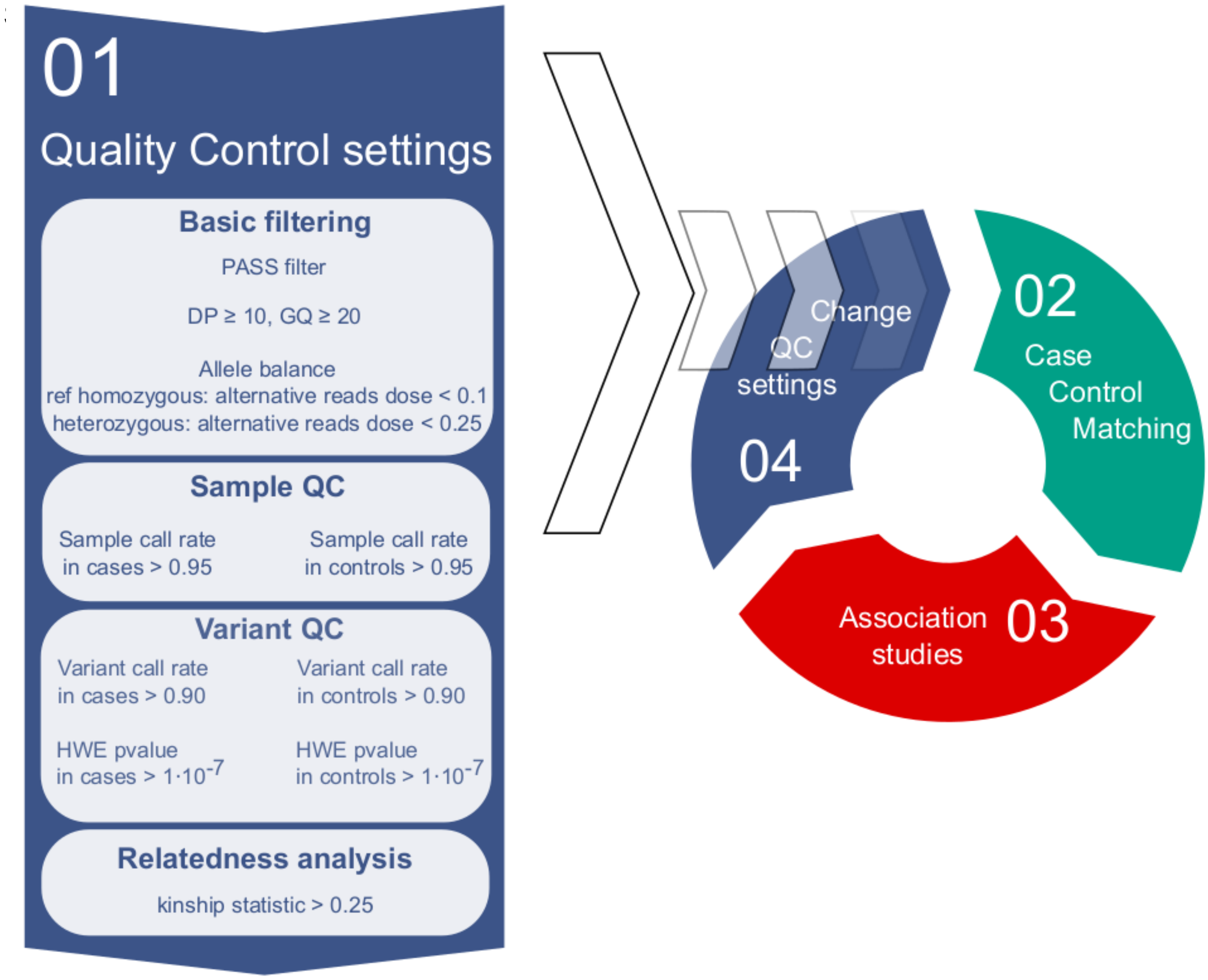


**Figure S2. Quality Control (QC) analysis. Filters threshold selection.**

The quality control was performed in Hail 0.2. Initially, we subjected raw genotypes to quality filtering, using DP>=10; GQ>=20 and an allele balance filter (0.25<AB<0.75 for heterozygotes and AB<=0.1 for alternative homozygotes). Only variants passing GATK filters ('PASS') were kept for further analysis.

We followed with sample and variant filtration using the Hardy-Weinberg equilibrium and call rate filters.

The final stage of QC is the relatedness analysis. Pairwise kinship was calculated using HAIL protocol: pc_relate(statistics='kin', min_kinship=0.25), maximum_independent_set(). For pairs with kinship > 0.25, only one of the samples was kept.

### Clustering on genotype data

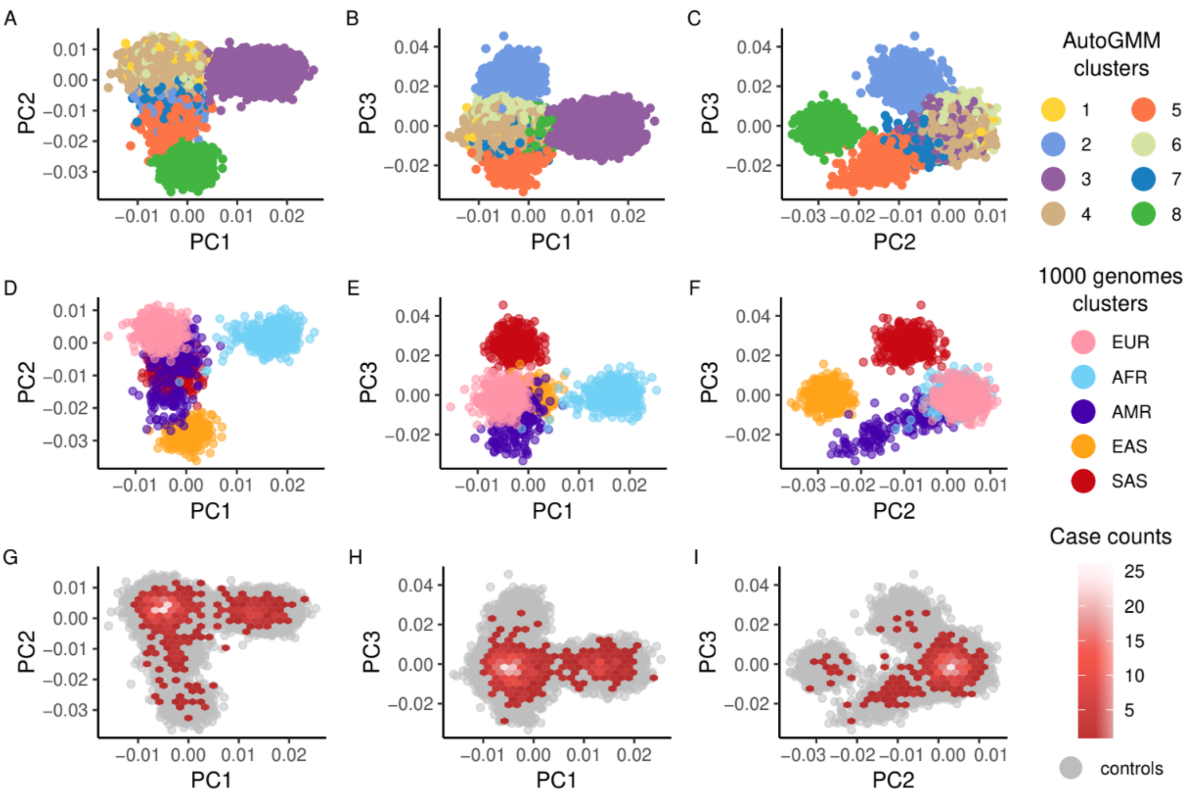


**Figure S3. Clustering using the AutoGMM mixed Gaussian model package.**

### Case-control matching

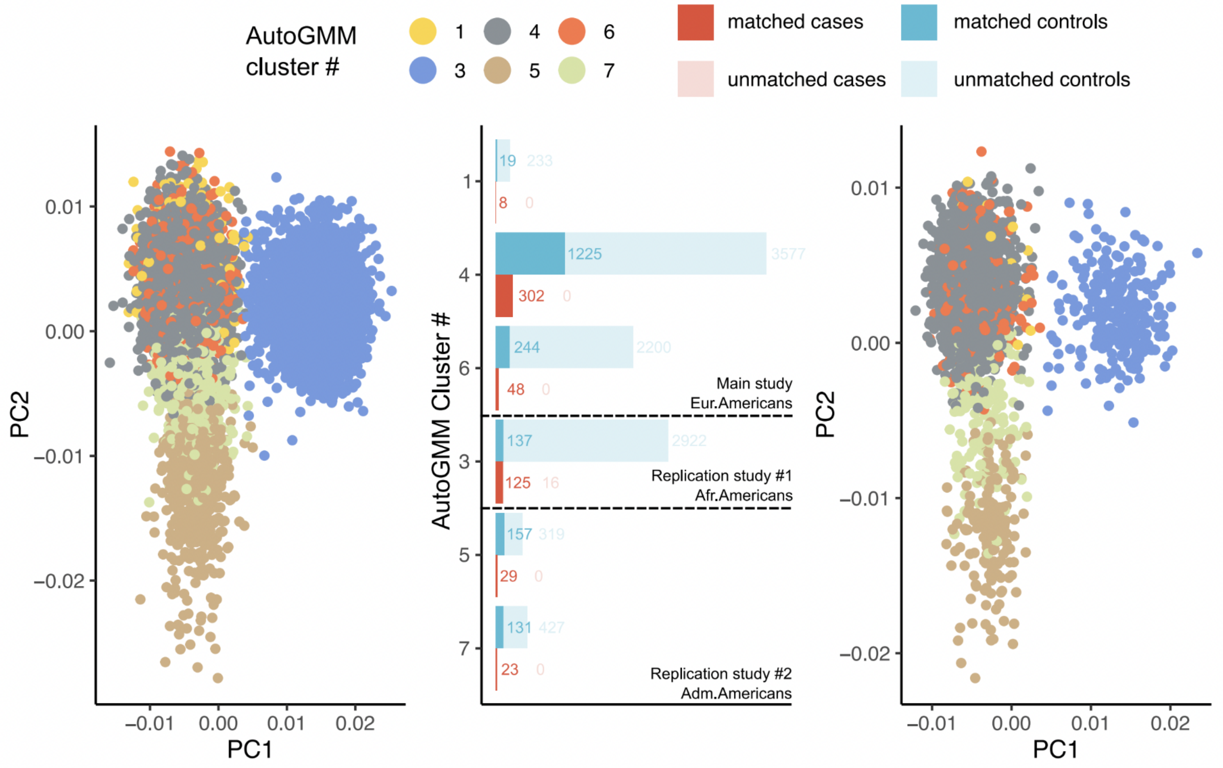


**Figure S4. Case-control matching. The left panel is colored according to the clusters assigned by the AutoGMM package before case-control matching. The middle panel shows the case-control composition of each cluster. The left panel is colored according to the clusters assigned by the AutoGMM package after case-control matching.**

To account for the effect of the population stratification component, the dataset passed QC was subjected to PCA and further clustering. Clustering was performed on 10 principal components using the mixed Gaussian model method using the AutoGMM package for python (AutoGMMCluster(affinity='euclidean')). The resulting clusters were named according to the continental populations and were validated by the presence of samples from 1000 genomes with the known ancestry.

#### Case-control matching

The cases belonging to individual European and African clusters were matched with controls from the respective clusters using the Matchit package for R. Matching was performed by downsampling the control dataset with respect to the principal components that contributed the most to the variance between the distributions of cases and controls.

#### Statistical power analysis

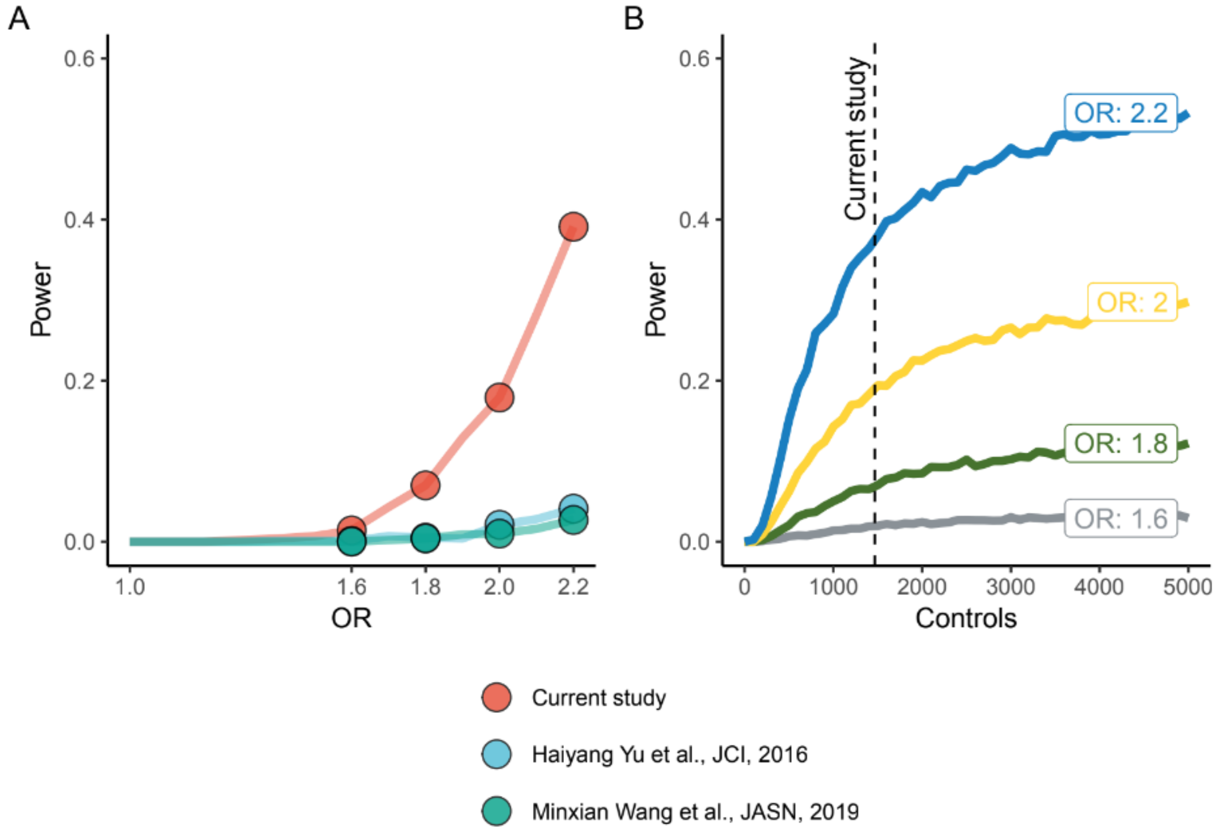


**Figure S5. Power analysis.**

Power analysis for the European cluster using matched cases and controls (for a hypothesized causal variant with MAF=0.05). We used next settings for the comparison (current study: 358 cases, 1488 controls, 2482 genes; Haiyang Yu et al, JCI, 2016: 179 cases, 378 controls, 2482 genes; Minxian Wang et al, JASN, 2019: 363 cases, 363 controls, 19000 genes). For the JASN study, we could not obtain the exact number of genes that were directly included in the analysis; therefore, the standard number of genes for whole-exome sequencing was used.

Alpha was calculated as a ratio of 0.05 to the number of genes used in a particular study ($a_{current\ study}$=0.05/2482, $a_{JCI}$=0.05/2482, $a_{JASN}$=0.05/19000).

We used the standard exact fisher test and self-written script in R for power estimation. Multiple odds ratios were used to demonstrate the dataset power for different types of risk variants

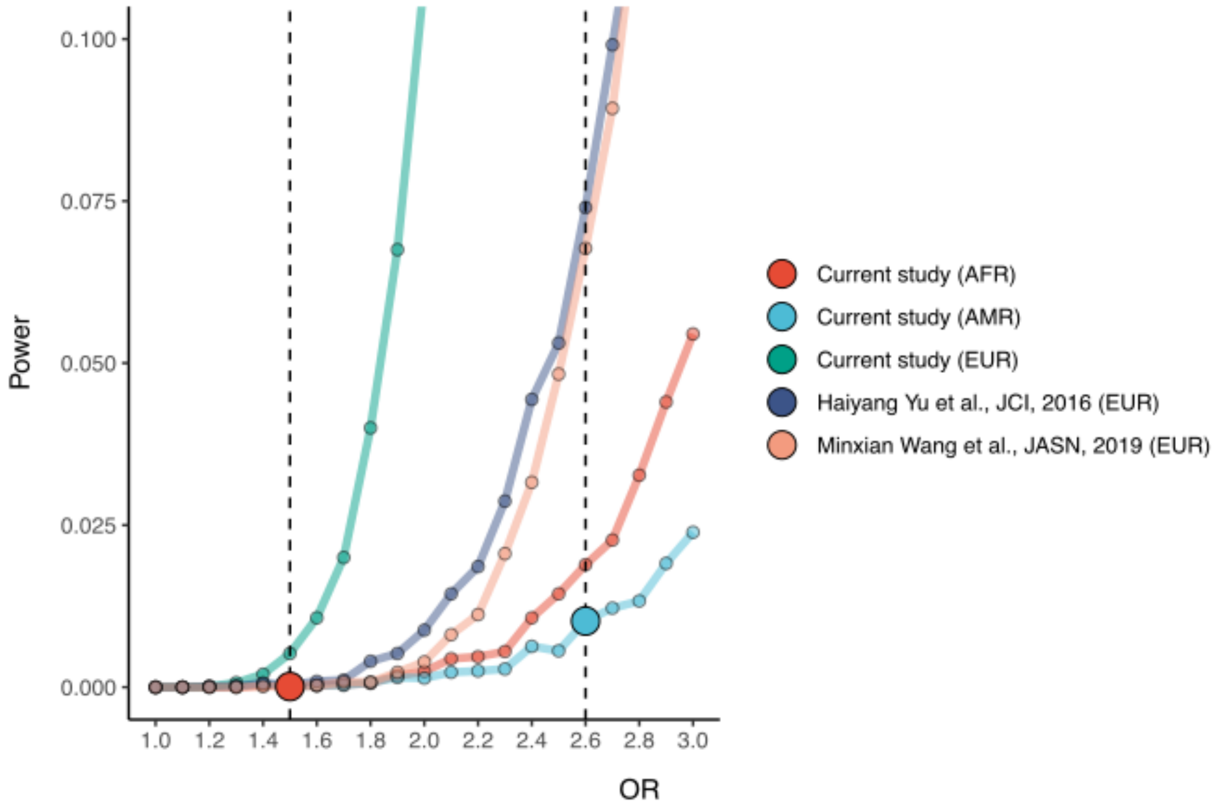


**Figure S6. Potential to detect statistically significant associations for a given effect size.**

### Association studies

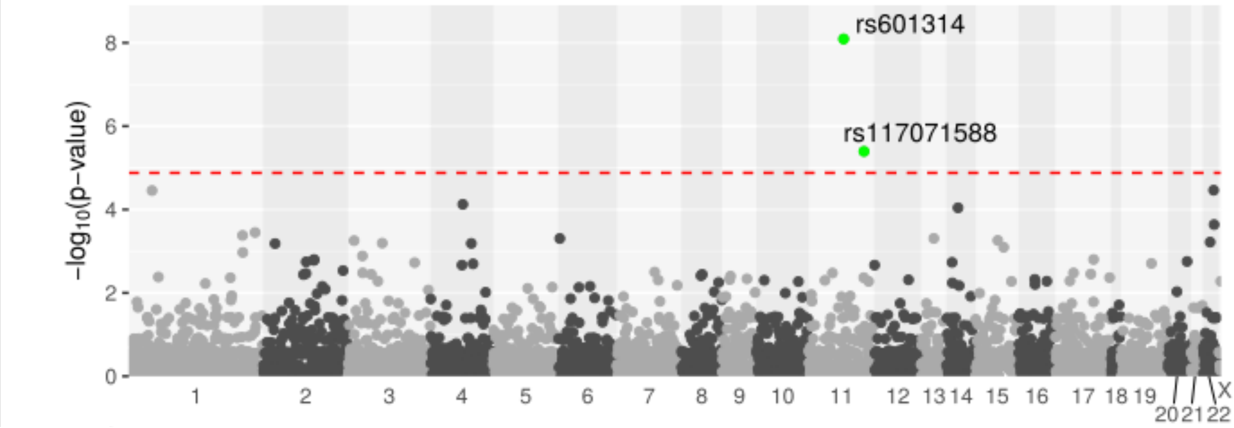


**Figure S7. Variant-based association analysis.**

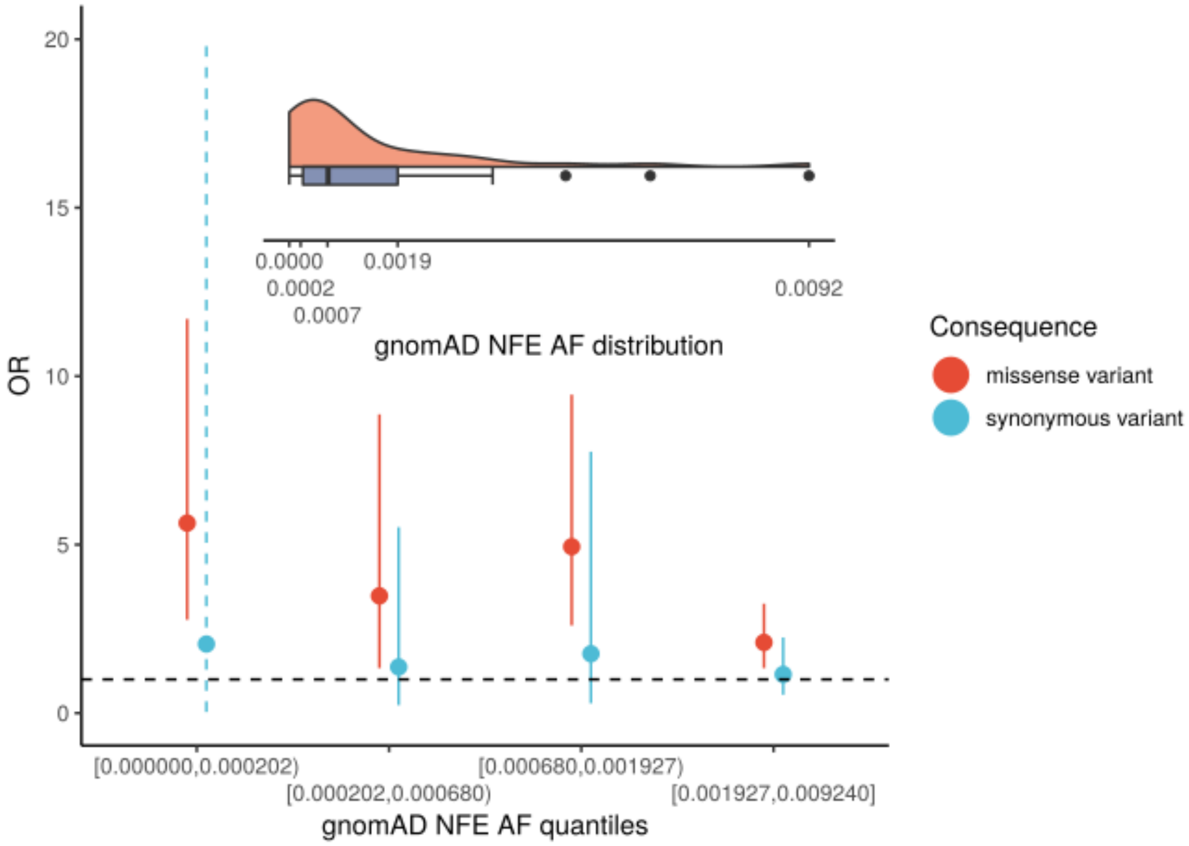


**Figure S8. Association signal strength among the known FSGS genes - *KANK1, COL4A4, WNK4, APOL1, IL36G* per gnomAD NFE allele frequency quantiles.**

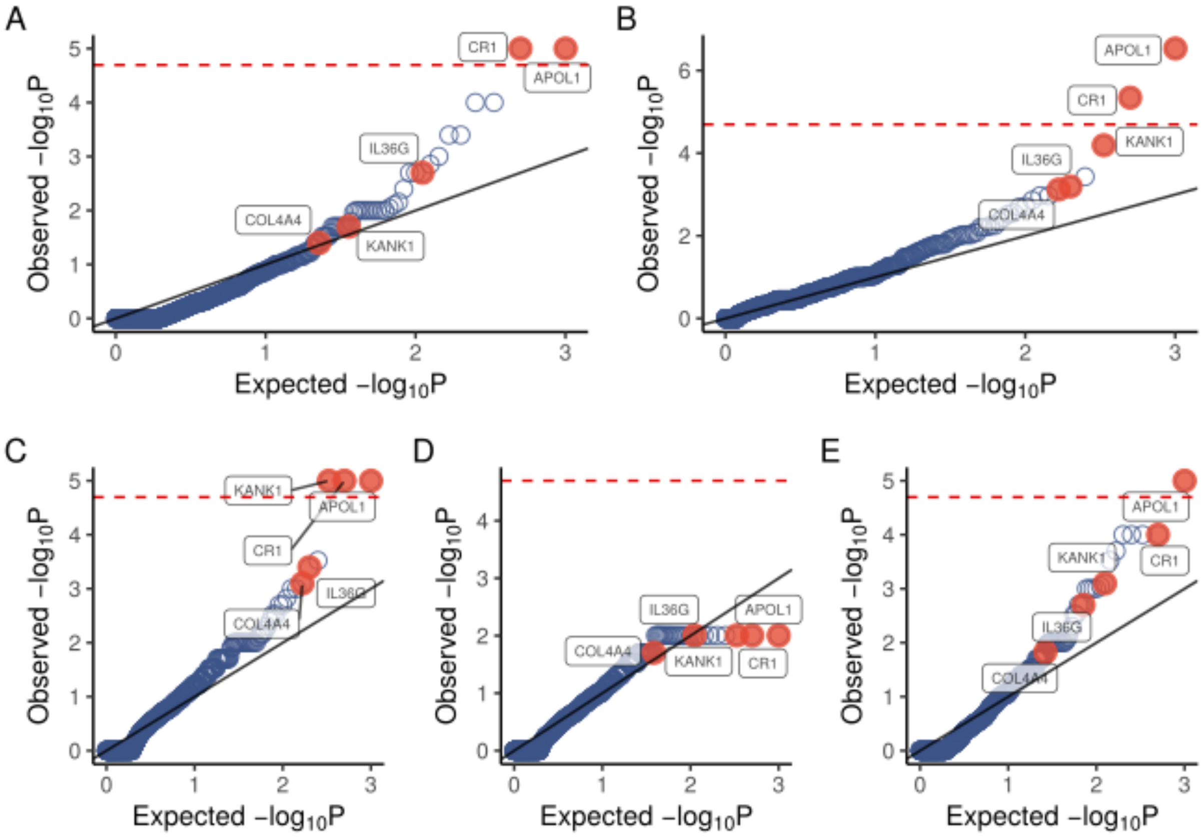


**Figure S9. QQ-plot for each of the rare variant tests included in the aggregating method of Simes. (A) C-alpha test; (B) Fisher's exact test; (C) WSS test; (D) KBAC test; (E) ASUM test**

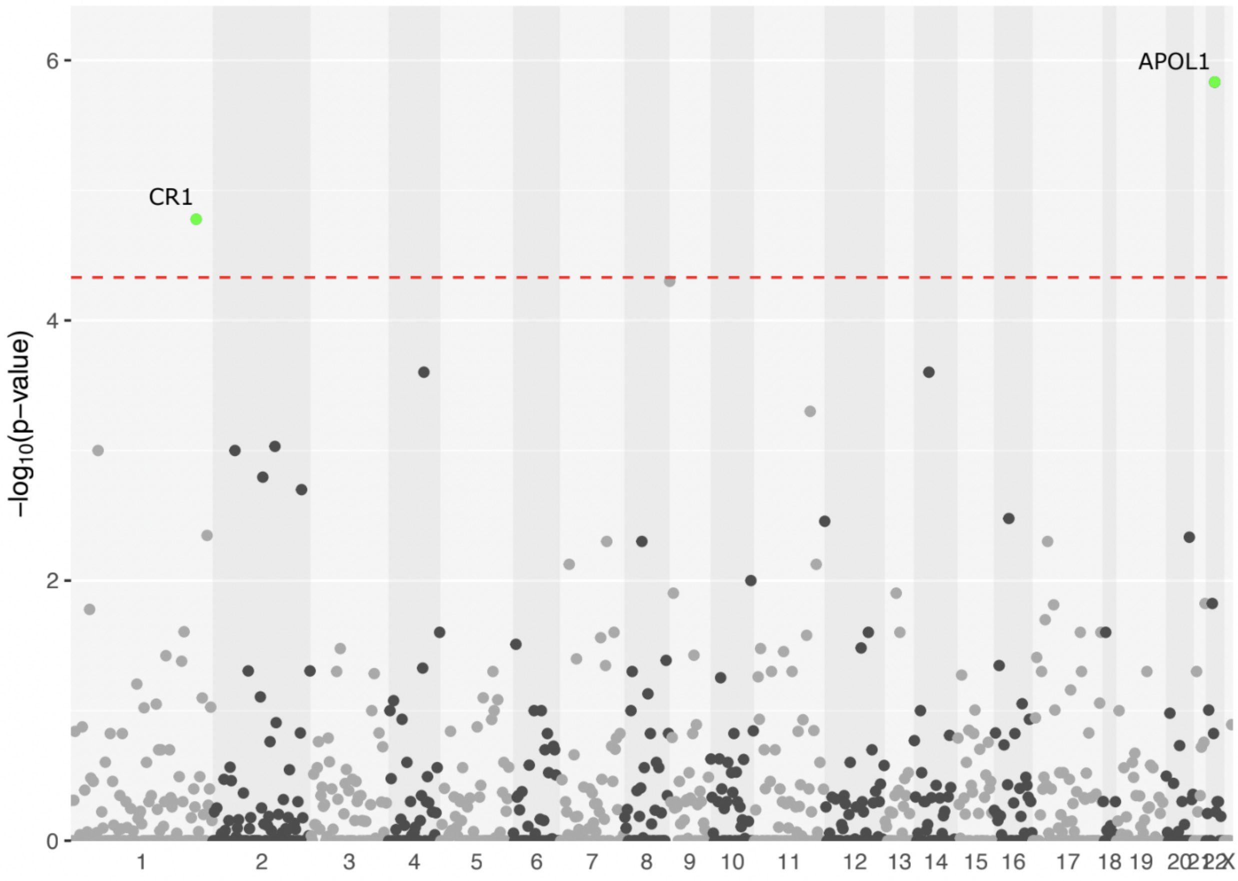

### Figure S10. Manhattan plot for RVAS in European-Americans

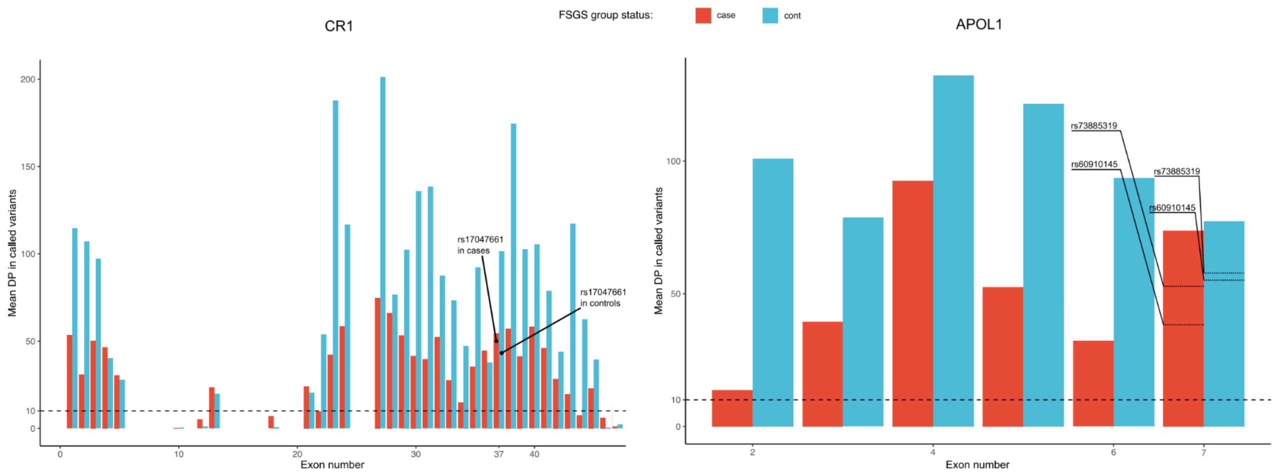


### Figure S11. Average coverage estimates of called variants for exons in cases and controls for *CR1* and *APOL1*.
